# Characterizing *SCN1A*-Related Disorders Using Real-World Data Across 681 Patient-Years

**DOI:** 10.64898/2026.02.24.26346493

**Authors:** Anna J. Prentice, Ian McSalley, Jan H. Magielski, Johanna Mercurio, Sarah Tefft, Angela Winters, Michael C. Kaufman, Sarah M. Ruggiero, Laura McGarry, Veronica Hood, Jillian L. McKee, Ethan Goldberg, Ingo Helbig

**Affiliations:** Division of Neurology, Children’s Hospital of Philadelphia, Philadelphia, PA, USA, 19104; The Epilepsy Neurogenetics Initiative (ENGIN), Children’s Hospital of Philadelphia, Philadelphia, PA, USA, 19104; Department of Biomedical and Health Informatics, Children’s Hospital of Philadelphia, Philadelphia, PA, USA, 19104; Center for Epilepsy and Neurodevelopmental Disorders (ENDD), Children’s Hospital of Philadelphia, Philadelphia, PA, USA, 19104; Dravet Syndrome Foundation, Cherry Hill, NJ, 08034; Department of Neurology, University of Pennsylvania Perelman School of Medicine, Philadelphia, PA, USA, 19104

**Keywords:** Epilepsy, genetics, neurogenetics, developmental and epileptic encephalopathy, Dravet syndrome, *SCN1A*

## Abstract

*SCN1A*-related disorders are the single most common monogenic cause of epilepsy and represent a major focus of precision medicine efforts. In conjunction with existing prospective studies, the analysis of real-world data obtained during routine clinical care can expand upon the scale and duration of available data and contribute to the development of meaningful outcomes for clinical trials.

Here, we leveraged real-world data to delineate the longitudinal disease history of 100 individuals with *SCN1A*-related disorders using a systematic approach. We mapped a total of 671 unique clinical terms to a standardized framework in monthly increments across 681 patient-years, including 75 terms related to seizure types. Within this cohort, 89 individuals had presumed loss-of-function variants in *SCN1A* based on variant type and clinical diagnosis, including those with Dravet syndrome (*N* = 79) and genetic epilepsy with febrile seizures plus (*N* = 10). Ten individuals had a non-Dravet developmental and epileptic encephalopathy caused by gain-of-function variants in *SCN1A*.

By annotating seizure type and frequency in monthly time-bins, we assessed seizure burden. A median of 17 changes in seizure frequency and ten terms referring to seizure type were identified per participant. Myoclonic seizures occurred with high frequency (median >5 daily), whereas hemiclonic, focal impaired consciousness, and bilateral tonic-clonic seizures occurred more rarely (median monthly). Retrospective analysis of developmental histories showed a range of cognitive abilities. Neurodevelopmental differences were observed in 83% (83/100) of individuals, of whom 83% (69/83) demonstrated delayed language skills. Motor coordination impairments, including gait disturbance, ataxia, hypotonia, and imbalance were annotated in 69% (69/100) of participants. EEG findings varied with age; most were reported as normal before nine months of age, after which the prevalence of abnormal interictal findings increased. Individuals with different clinical syndromes had unique medication landscapes, with 554 prescriptions of 37 unique therapies. Changes in treatment coincided with the diagnosis of an *SCN1A*-related disorder, with an increase in cannabidiol, clobazam, and fenfluramine and reduction in sodium channel–blocker use following genetic diagnosis.

In summary, we reconstructed the longitudinal disease history of *SCN1A*-related disorders from electronic medical records using a standardized framework for the analysis of real-world clinical data. We refine existing natural history data of *SCN1A*-related disorders by providing a granular landscape of seizures, comorbidities, and treatment approaches over time.

## Introduction

Disease-causing variants in *SCN1A* cause one of the most clinically recognizable and well-studied monogenic epilepsies with a prevalence of 1 in 15,700.^1^ A spectrum of disorders are associated with *SCN1A*, the most notable of which are genetic epilepsy with febrile seizures plus^2^ (GEFS+) and Dravet syndrome.^3^ GEFS+ was first characterized in large kindreds of individuals with fever-sensitive seizures, often recurring beyond the typical age range of febrile seizures and followed by unprovoked seizures.^4^ Epilepsy in Dravet syndrome onsets in the first year of life—most frequently four to six months of age—in a previously typically-developing infant, often consisting of prolonged, alternating hemiclonic seizures. Later, seizures may be provoked or unprovoked and include bilateral tonic-clonic (BTC), myoclonic, atypical absence, and focal semiologies. Seizure precipitants may evolve with age, and include fever, heat, stress, exercise, sleep deprivation, and visual/auditory stimuli.^5^ Epilepsy in Dravet syndrome is often refractory and is a major target of therapy development. Unlike GEFS+, Dravet syndrome has prominent neurodevelopmental co-morbidities, including hypotonia, ataxia, and developmental delay.^6^ 90% of cases of Dravet syndrome are the result of *de novo* variants in *SCN1A*, with a small percentage attributed to parental mosaicism.^7,8^

The spectrum of *SCN1A*-related disorders also includes gain-of-function (GoF) variants, which can cause an early infantile-onset disorder with features distinct from both GEFS+ and Dravet syndrome. Existing literature refers to this disorder by several names, including neonatal developmental and epileptic encephalopathy (DEE) with movement disorder and arthrogryposis (NDEEMA) and early infantile DEE (EIDEE).^9^ Formal curation of *SCN1A* by the Epilepsy Gene Curation Expert Panel selected DEE as the gene-disease relationship. Given that not all individuals with this phenotype present with a movement disorder, arthrogryposis, or early infantile seizures, and to distinguish it from Dravet syndrome, we will refer to it as non-Dravet syndrome DEE (nd-DEE). For these individuals, seizures often onset before three months of age and are rarely provoked. Tonic seizures, unusual in GEFS+ and Dravet syndrome, are frequent. Similarly to Dravet syndrome, BTC, focal, and myoclonic seizures are common. Contractures and hyperkinetic movement disorders also distinguish this disorder from Dravet syndrome and GEFS+.^10^ Whole-cell voltage clamp studies, have demonstrated mixed effects with an overall gain of function, including a hyperpolarized shift of steady-state inactivation and a persistent sodium current.^10^

At the molecular level, *SCN1A* encodes the alpha subunit of the voltage-gated sodium channel NaV1.1, expressed predominantly in GABAergic inhibitory interneurons.^11^ Haploinsufficiency of this channel, caused by loss-of-function (LoF) variation in *SCN1A*, causes Dravet syndrome and GEFS+, whereas GoF variation causes the nd-DEE phenotype.^10^ The later onset of disease associated with LoF variants is presumed to be caused by a developmental switch. During early development, most *SCN1A*-encoded mRNA results in a nonfunctional protein product due to the inclusion of a naturally occurring poison exon. Later, this exon is skipped, and functional protein is made.^12,13^ *SCN1A* haploinsufficiency is therefore typical during early development but becomes pathologic with age. Alternatively, channel dysregulation caused by GoF variants is disruptive at all developmental stages, resulting in the earlier onset of disease.

Though much is known about the natural history of *SCN1A*-related disorders and their underlying pathomechanisms, a month-by-month analysis of seizures and comorbidities leveraging a standardized phenotypic vocabulary has not yet been performed. Natural history studies often employ differing methodologies, such as investigator-created surveys with pre-defined symptoms of interest reported at varying levels of detail. This renders comparisons between studies and across diseases difficult. Data collected clinically is similarly nonstandard. Vast amounts of information are available in the electronic medical record (EMR) of individuals with *SCN1A*-related disorders, though they are typically written in a form difficult to leverage for large-scale analysis. Efforts to use this information include translation into terms from a standardized biomedical dictionary. Resources such as the Human Phenotype Ontology (HPO) are now used in both clinical and research settings to communicate patient phenotypes in a standardized manner. The ontological structure of the HPO—wherein terms are related hierarchically—enables the systematic comparison of patient phenotypes both within and between genetic disorders.^14–17^

In addition to information collected in clinical trials, a systematic assessment of the natural history of *SCN1A*-related disorders using real-world data is a key step toward clinical trial readiness, particularly for a disease entity with multiple ongoing trials for targeted therapeutics. Not only can retrospective studies incorporate a larger study population for minimal cost, but they also permit the use of data obtained prior to a participant’s genetic diagnosis and enable the inclusion of individuals who would not otherwise be able to participate in a prospective study based on location or time required. Here, we performed a retrospective phenotypic analysis of 100 individuals to create a detailed phenotypic landscape across the lifespan of individuals with *SCN1A*-related disorders in a standardized language amenable to future comparisons.

## Materials and methods

### Cohort Identification

Our cohort consisted of 100 individuals with *SCN1A*-related disorders recruited at a tertiary care center and throughout the United States via the Dravet Syndrome Foundation. This sample size is comparable to similar studies of rare genetic epilepsies.^16^ Clinical documentation was gathered for each patient. Records were reviewed for the participant’s entire lifespan from birth until the date of review, which occurred from 2023–2024. *SCN1A* variants were converted into standard HGVS nomenclature. Clinical Genome Resource (ClinGen) <u>Epilepsy Sodium Channel Variant Curation Expert Panel</u> (ESC-VCEP) guidelines were used to curate each *SCN1A* variant to confirm genetic diagnosis and study eligibility. Evidence criteria applied to each variant are available in **Supplementary Table 2**.

### Clinical Data Review and Standardization

EMR data was reviewed manually for each participant. Phenotypes were translated from free text into HPO terms. Age in months was recorded for each term based on the date of EMR entry. Epilepsy phenotypes were assigned a frequency score using a modified scale endorsed by the Epilepsy Foundation’s Epilepsy Learning Health System and Pediatric Epilepsy Learning Healthcare System.^18^ Scores range from 0–5 (0 = none, 1 = monthly, 2 = weekly, 3 = daily, 4 = 2–5 daily, 5 = >5 daily). In this way, seizure semiologies and their frequencies were reconstructed monthly at all ages where data was available. Missing data was assigned N/A and not included in analysis. Basic demographic and clinical information are reported in **Supplementary Table 1**.

EEG reports and treatment data were also extracted from the EMR. EEG reports were assessed for HPO terms as described above. Age in months and normal/abnormal status was recorded for each EEG. Clinical notes were reviewed for therapy use. The month of age of initiation and discontinuation of each treatment was recorded.

After extracting phenotypic data, HPO terms were propagated, meaning all higher-level terms in the ontological structure were assumed to be present. Seizure frequencies were propagated as well, assigning the highest frequency among more specific terms to their common ancestor. This methodology has been employed previously.^14–16,19^

### Statistical Analysis

Statistical analyses were conducted using the R statistical framework.^20^ The frequency of each phenotypic term was calculated across the entire cohort and across subgroups. Fisher Exact and Wilcoxon Rank-Sum tests were used to assess clinical feature associations, compare symptom onset between subgroups, and assess the comparative effectiveness of epilepsy treatments as described previously.^16^

### Ethics and Regulatory Approval

Participants or their guardians provided informed consent according to the Declaration of Helsinki. This study was approved by the Institutional Review Board of the Children’s Hospital of Philadelphia (IRB 15-012226).

## Results

### Individuals with *SCN1A*-related disorders are clinically and genetically diverse

The final dataset included clinical data from 100 individuals in 95 families with *SCN1A*-related disorders, assembled agnostic of clinical diagnosis. Participant ages ranged from seven months to 39 years (median seven years). These individuals were followed by 50 neurology providers, including 46 epileptologists or neurologists (93/100 participants) and four advanced practice providers (7/100 participants). Clinical diagnoses were recorded based on the diagnostic labels assigned by providers during clinical care.

Of the 100 participants, 79 were diagnosed by their provider with Dravet syndrome, of whom 84% (67/79) were considered to have typical Dravet syndrome. The remaining 16% (13/79) were assigned clinical diagnoses of atypical Dravet syndrome, a phenotypic category formerly referred to as severe myoclonic epilepsy of infancy borderline (SMEB).^21^ Ten individuals were diagnosed with GEFS+ and ten with nd-DEE. One participant displayed an epilepsy phenotype that was not clearly consistent with the above categories and had a clinical diagnosis of Lennox-Gastaut syndrome. Demographic and clinical information for each participant is reported in **Supplementary Table 1**.

It is well established that Dravet Syndrome and GEFS+ are caused by LoF variants in *SCN1A*.^22,23^ Additionally, recent literature suggests that nd-DEE is the result of GoF variants.^9,10^ Accordingly, the 90 individuals diagnosed with either Dravet syndrome, GEFS+, or an unclassified phenotype had presumed LoF (pLoF) variants in *SCN1A*. Of these 90 participants, 47% (42/90) had predicted null variants, including frameshift, nonsense, and splice site variants as well as multi-exon duplications or deletions. 49% (44/90) had missense variants, three had intronic variants, and one had an in-frame deletion. 79% (50/63) of pLoF variants with available inheritance information occurred *de novo*. Dravet syndrome was more likely to be caused by a *de novo* variant than GEFS+ (48/52 Dravet; 2/8 GEFS+; *P* < 0.001). Recurrent variants were rare in the pLoF cohort: 85% (73/86) of probands had private variants. Only one variant was present in more than 5% of individuals: p.Lys1846SerfsTer11, which was observed in six unrelated participants.

Of the ten individuals with the nd-DEE phenotype, seven had the established GoF variants p.Arg1636Gln (*N* = 4) and p.Ile1347Val (*N* = 3). The three remaining individuals harbored p.Leu209Arg, p.Ser872Tyr, and p.Ala889Thr, which have not previously been reported in association with nd-DEE. However, for each of these three variants, paralogous variants have been observed in *SCN2A*, *SCN3A*, or *SCN8A* in patients with clinical presentations consistent with GoF (*SCN2A* p.Ala880Thr, p.Ser863Tyr; *SCN3A* p.Leu209Pro; *SCN8A* p.Ala874Thr).^24–27^ Paralogous variation across sodium channel–encoding genes is emerging as a means to provide evidence of variant pathogenicity and demonstrates functional consistency across a gene class.^10,28^ Therefore, when combining clinical and variant-level evidence, all ten individuals with nd-DEE were presumed to harbor gain-of-function (pGoF) variants in *SCN1A*. Across the cohort, variants were located throughout the entire *SCN1A*-encoded NaV1.1 protein. Missense variants were more frequently found in critical transmembrane or pore-forming domains when compared to predicted null variants (*P* < 0.001).

### A quarter of diagnostic *SCN1A* findings are variants of uncertain significance

Despite increased emphasis on genetic testing, Dravet Syndrome and GEFS+ remain clinical diagnoses. Accordingly, our real-world dataset allowed us to contrast clinical diagnosis with variant evidence. In clinical practice, variants lacking sufficient evidence of pathogenicity are labelled variants of uncertain significance (VUS). When VUS are presumed to cause a patient’s symptoms, they are referred to as “explanatory” or “diagnostic.” Given that variant classification criteria were updated by the ClinGen ESC-VCEP in 2025, we assessed the evidence supporting each variant identified in this cohort.^29^ Copy number variants were assessed as outlined by the joint ClinGen/ACMG/AMP consensus recommendations.^30^

Though all variants were considered diagnostic, only 76% (61/80) of variants met criteria to be classified as pathogenic or likely pathogenic. 24% (19/80) of variants, including 41% (17/41) of missense variants, were therefore classified as VUS. 97% (75/77) of variants (excluding copy number variants) were present ≤1 time in population databases (gnomAD v4.1.0), providing supporting evidence of pathogenicity. Of variants where *in silico* predictors were available, 38/39 were deemed likely deleterious, contributing supporting (3) or moderate (35) evidence. Additional evidence was applied for each proband and for cases reported in the literature as observations of the variant in affected individuals. The full list of criteria met for each variant is reported in **Supplementary Table 2**. In summary, the spectrum of *SCN1A*-related disorders encompasses at least three clinical syndromes, two molecular mechanisms, and a range of established and emerging variants, contributing to a diverse and variable disease landscape.

### Features of *SCN1A*-related disorders can be captured by longitudinal disease reconstruction

While prospective natural history studies are the gold standard to resolve disease trajectory and develop outcomes for clinical trials, the scalability of this approach is limited. Accordingly, recent efforts have focused on repurposing real-world clinical data to elucidate the natural history of genetic epilepsies, including those caused by *STXBP1*, *SYNGAP1*, and *SCN8A*.^15,16,31^ Here, we used a standardized biomedical ontology to transform natural language data from medical records into a standardized format. 8,171 patient-months of data were reviewed for the 100 participants. Phenotypes were recorded in one-month time bins, resulting in 671 unique clinical terms mapped to 2,220 distinct observations across the cohort.

A key feature of biomedical ontologies is their hierarchical structure, where terms more general than those assigned manually can be inferred. For example, if an individual is assigned the phenotype “Myoclonic seizure,” (HP:0032794) term propagation would assign both “Motor seizure” (HP:0020219) and “Seizure” (HP:0001250). Term propagation expanded the dataset to 1,174 unique features assigned 9,176 times. Mean phenotypic depth increased concomitantly—from 22 to 92 terms per individual. Of all terms, 75 referred to aspects of seizure semiology and 1,103 referred to non-seizure phenotypes, including developmental differences, behavioral abnormalities, and a range of additional symptoms. Well-recognized clinical features of *SCN1A*-related epilepsies were observed frequently, demonstrating that our method of disease reconstruction recapitulated known aspects of *SCN1A*-related disorders (**Fig. 1A**). Overall, the longitudinal phenotypic profiling of 100 individuals with *SCN1A*-related epilepsies yielded a dataset encompassing 681 patient-years and 9,176 clinical annotations, reconstructing the natural history of these disorders in a complementary manner to prospective studies.

**Figure 1.**
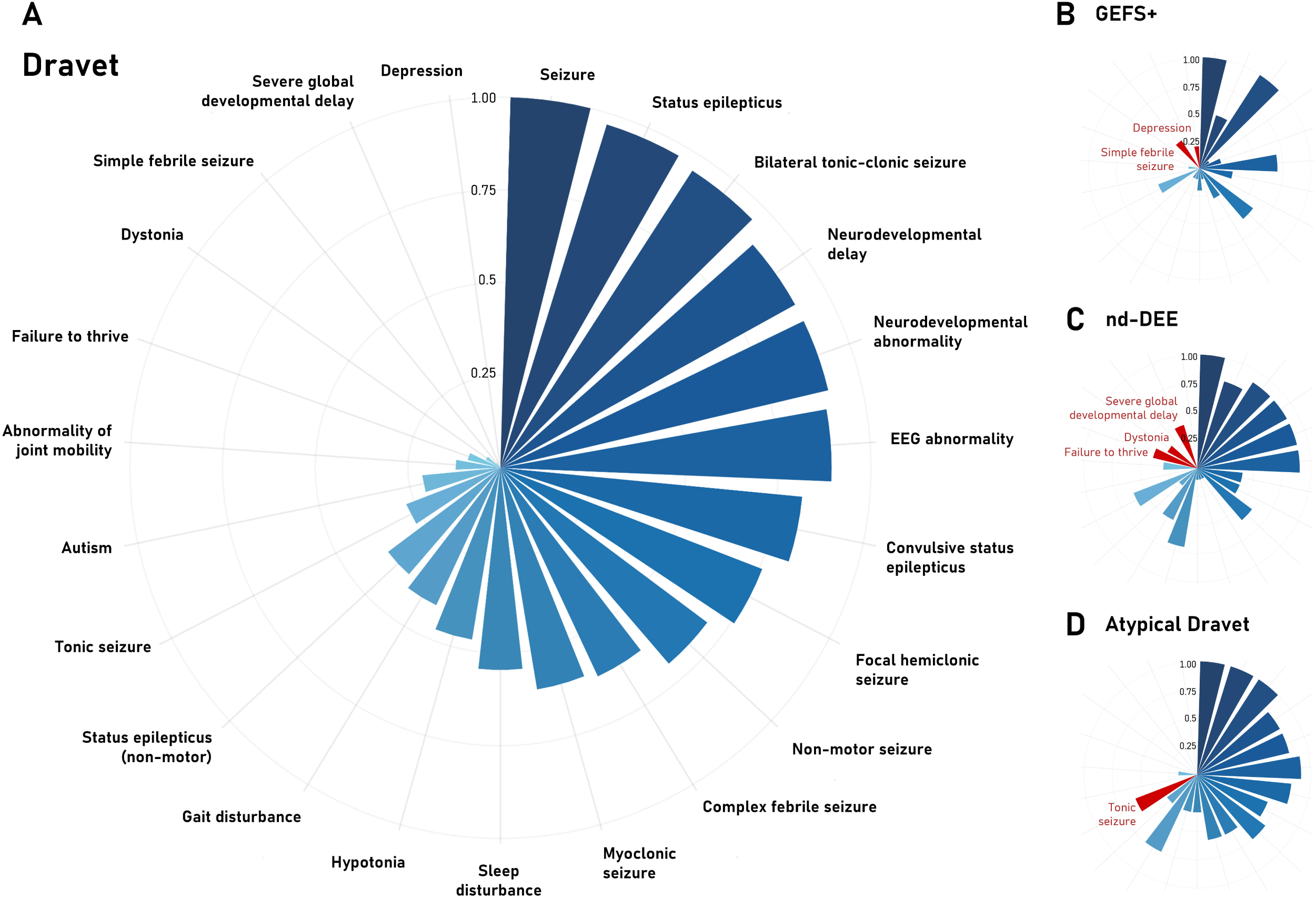
Frequency of select phenotypic features by clinical diagnosis. The frequency of selected phenotypes in individuals with Dravet syndrome (**A**), genetic epilepsy with febrile seizures plus (GEFS+; **B**), non-Dravet developmental and epileptic encephalopathy (nd-DEE; **C**), and atypical Dravet syndrome (**D**). Phenotypes are sorted by frequency in Dravet syndrome. Terms highlighted in red are enriched compared to the Dravet syndrome sub-cohort at *P* < 0.05.

### Diagnostic variants in *SCN1A* cause distinct clinical syndromes

The distinct clinical syndromes caused by variation in *SCN1A* are united by the presence of multiple seizure types in all individuals, demonstrating a high burden of epilepsy. BTC seizures (96/100) and status epilepticus (96/100) stood out as near-universal features, followed by seizures precipitated by febrile infection (81/100), focal motor seizures (72/100), convulsive status epilepticus (69/100), and focal hemiclonic seizures (64/100; **Fig. 1A**), seizure types known to be associated with *SCN1A*-related disorders. Reflective of this high seizure burden, 76/100 individuals had at least one episode of respiratory desaturation associated with a seizure and 57/100 had at least one seizure cluster. Despite the initial identification of Dravet syndrome as severe myoclonic epilepsy of infancy, only 41/79 individuals with Dravet syndrome had myoclonic seizures.

We then compared phenotypic terms to clinical diagnosis. Though some features were common across clinical diagnoses, others distinguished them. Compared to individuals with Dravet syndrome, individuals with GEFS+ were less likely to have neurodevelopmental delay (*P* < 0.001), status epilepticus (*P* < 0.001), myoclonic seizures (*P* = 0.004), hypotonia (*P* = 0.038), or hemiclonic seizures (*P* < 0.001) (**Fig. 1B)**. Conversely, these participants were more likely to have simple febrile seizures (*P* = 0.006) and depression (*P* = 0.016). The milder disease course of GEFS+ was also reflected in the number of terms assigned. Those with GEFS+ had roughly half the number of clinical annotations as those with Dravet syndrome (mean 54 vs. 98 terms per individual; *P* < 0.001). This pattern remained consistent when restricted to seizure phenotypes (mean 14 vs. 22 terms per individual; *P* < 0.001). Participants with the nd-DEE phenotype had a unique phenotypic landscape (**Fig. 1C**). Symptoms enriched in this sub-cohort compared to Dravet syndrome included joint contractures (*P* = 0.006), severe global developmental delay (*P* < 0.001), abnormal brain morphology (*P* = 0.002), dystonia (*P* = 0.027), and failure to thrive (*P* = 0.023). These individuals were less likely to have seizures triggered by fever (*P* = 0.003), myoclonic seizures (*P* = 0.004), or convulsive status epilepticus (*P* = 0.009). Diagnoses of classical and atypical Dravet syndrome, as assigned by providers, were difficult to disentangle. Only six clinical terms were either enriched or depleted in those with an atypical presentation, including a higher frequency of tonic seizures (*P* = 0.046) and cardiac arrhythmia (*P* = 0.028; **Fig. 1D**). In summary, the clinical diagnoses associated with *SCN1A* are distinguished by clinical features, including the number of terms assigned and the presence or absence of certain symptoms. Classical and atypical Dravet syndrome displayed minimal differences, supporting emerging evidence that they may not be distinct entities.^21^

### Highly variable seizure burdens are seen in individuals with *SCN1A*-related disorders

We next focused on the longitudinal trajectory of seizures among participants. Although all individuals had seizures, onset and semiology varied widely (**Fig. 2A**). For individuals with Dravet syndrome, seizure onset occurred from 1–22 months, with a median of five months. Onset was later for GEFS+ (14 months, *P* < 0.001). Of those with the nd-DEE phenotype, 8/10 had seizure onset from 0–4 months of age. The other 2/10 were identical twins whose first definite seizures occurred at 11 months, though they displayed abnormal movements in early infancy that were not confirmed to be epileptic. Taken together, this sub-cohort had an earlier seizure onset (median 2.5 months, *P* = 0.014) than individuals with Dravet syndrome (**Fig. 2B**).

**Figure 2.**
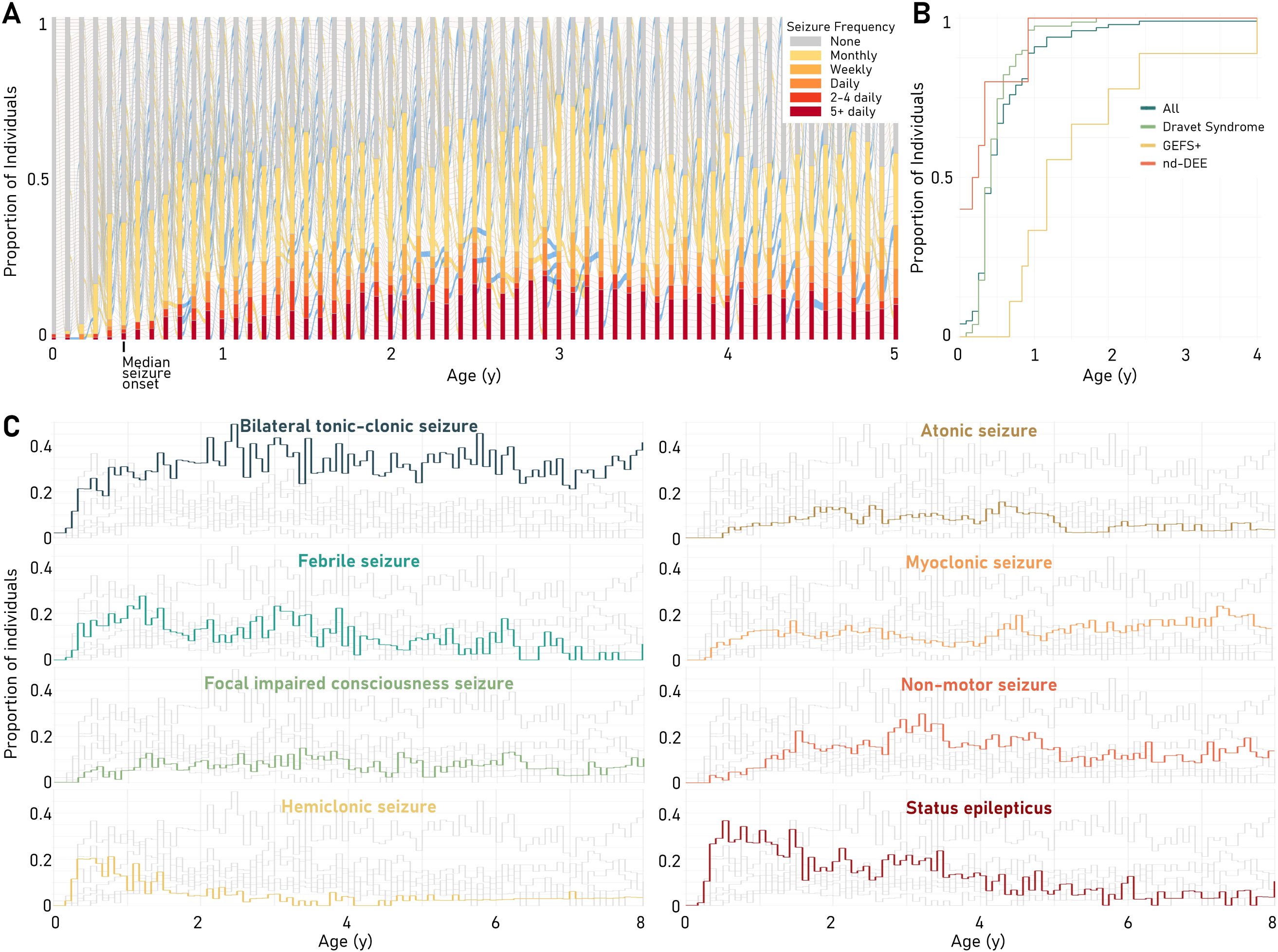
**Seizure frequency across the lifespan**. (**A**) Seizure frequency over the first five years of life. Each vertical bar represents one month of life. Bar color represents the proportion of individuals with a given seizure frequency as described in the legend. Blue and yellow connections between bars represent a decrease or increase in seizure frequency between months, respectively. (**B**) Cumulative seizure onset by clinical diagnosis. Y-values represent the proportion of individuals who have had least one seizure by the age indicated on the x-axis. (**C**) Seizure semiologies over the first eight years of life. One seizure type is highlighted in color per panel, with others shown in grey. At each age, line height represents the proportion of individuals for whom data is available having the seizure type indicated.

Across the cohort, seizure onset occurred within a narrow window. Between three and five months, incidence of seizures increased from <20% to >50% of individuals. The proportion of individuals with active epilepsy, defined as a seizure frequency greater than zero, remained at or above 50% between one and five years of age, peaking at 38 months when over 75% of individuals had seizures.

Semiology varied with aged. Early life was characterized by status epilepticus and hemiclonic seizures, which affected the highest proportions of individuals before one year of age (37% and 22% monthly incidence, respectively). Seizures triggered by fever were also common at early ages with an overall prevalence of 82% and a monthly incidence peaking at 28% at 14 months. After these early peaks, all three seizure types became less prominent. Alternatively, BTC seizures occurred frequently both in infancy and childhood. Between one and eight years of age, 21–49% (median 32%) of individuals had BTC seizures each month. Myoclonic seizures displayed a similar pattern, though they affected fewer participants (5–24% per month, median 12%). Atonic seizures, non-motor seizures, and focal impaired consciousness seizures affected smaller proportions of participants and often onset later in childhood, (median 6%, 4%, and 7% per month, respectively from age 1-8; **Fig. 2C**). Overall, our reconstruction of real-world data shows that childhood with an *SCN1A*-related disorder is characterized by persistent seizures despite treatment, though myoclonic seizures were not observed in most individuals.

### Seizure frequency in *SCN1A*-related epilepsies is multidimensional

We next focused on seizure frequency across the cohort. Of those with Dravet syndrome, a range of frequencies were observed, from seizure freedom to many daily seizures. At all months before age five, at least one individual was seizure-free and one had daily seizures, highlighting the large variability in clinical presentations. Of those with seizures (frequency > 0) in a given month, the most common frequency was monthly, followed by weekly and then > 5 daily (**Fig. 2A**). 76/100 individuals had monthly seizures for more than six months during their lifetime, whereas only 45/100 had weekly or more frequent seizures for this length of time. All participants with GEFS+ were seizure-free for more than half the months surveyed. Seizure frequency was significantly lower than in Dravet syndrome (*P* < 0.001). For nd-DEE, though the temporal dynamics of seizures differed from Dravet syndrome, overall seizure frequency did not differ.

Seizure frequency also varied month-to-month. The average individual had a median of three seizure frequencies and 17 alterations in seizure frequency throughout their life (**Fig. 2A, 3A**). Frequencies were not distributed equally by seizure type. Fever-triggered seizures, status epilepticus, and BTC seizures occurred almost exclusively monthly, whereas myoclonic seizures often occurred daily. Semiologies of intermediate frequency included hemiclonic seizures and focal impaired consciousness seizures, which tended to occur weekly or monthly (**Fig. 3B-D**).

**Figure 3.**
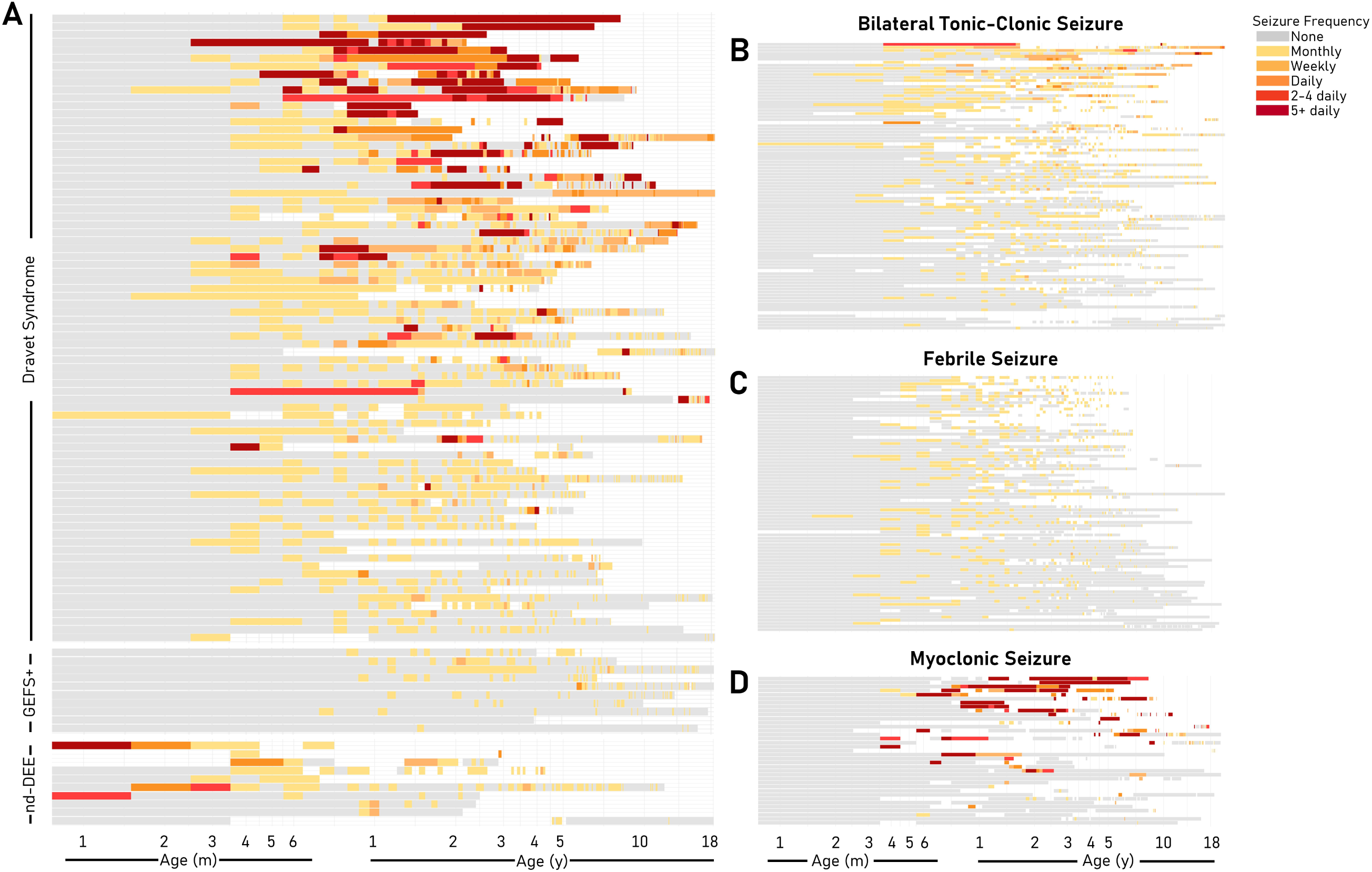
**Seizure frequency by individual and semiology**. Seizure frequency is represented overall (**A**) and by semiology (**B-D**) over the first 18 years of life. Each segment of the y-axis represents one individual, with panel A being grouped by clinical diagnosis: Dravet syndrome (top), genetic epilepsy with febrile seizures plus (GEFS+; middle), and non-Dravet developmental and epileptic encephalopathy (nd-DEE; bottom). The x-axis represents age on a logarithmic scale from 0-18 years. Seizure frequency is indicated by color as outlined in the legend. White space indicates months where seizure frequency is unknown for a given individual.

### Behavior, development, and motor differences are key features of *SCN1A*-related disorders

Neurodevelopmental and behavioral phenotypes commonly reported in association with Dravet syndrome were also observed in this cohort. Behavioral disturbances, including maladaptive behavior (45/100), sleep disturbance (43/100), and attention difficulties (22/100), were reported in 68/100 individuals (**Fig. 4A**). Only 14/100 individuals carried a formal diagnosis of autism spectrum disorder, which is much lower than that reported in studies assessing rates of autism in Dravet syndrome using standardized tools (24–40%).^32^ The proportion of participants reported to exhibit autistic features, however, falls within this range (28/100). Aggressive behavior was observed in 25/100 participants. Most behavioral features onset between one and ten years of age, though attention difficulties were often first reported between 3–6 years of age (14/22). Cognitive differences affected 83/100 individuals (**Fig. 4B**). The most commonly impaired domain was speech and language (69/100). Delayed motor development was less common, affecting 31/100 individuals, including gross motor (20/100) and fine motor (16/100) delays. Despite the high rate of developmental delay, only 19% of those over five years of age had a diagnosis of intellectual disability. This value—obtained from real-world data as opposed to formal developmental assessments—is lower than would be suggested by findings of prospective natural history studies of Dravet syndrome, which suggest plateauing around the developmental age of two years.^33^ Although motor delays affected only one-third of participants, 69/100 individuals went on to develop impaired motor function (**Fig. 4C**). This category of symptoms included abnormal tone (42/100), incoordination (43/100), gait disturbance (41/100), and imbalance (33/100). Of these features, hypotonia was observed at the earliest ages, with a median onset of two years. Gait disturbances were characterized by broad-based gait (20/100), ataxia (10/100), unsteadiness (7/100), tiptoeing (6/100), and crouch gait (5/100).

**Figure 4.**
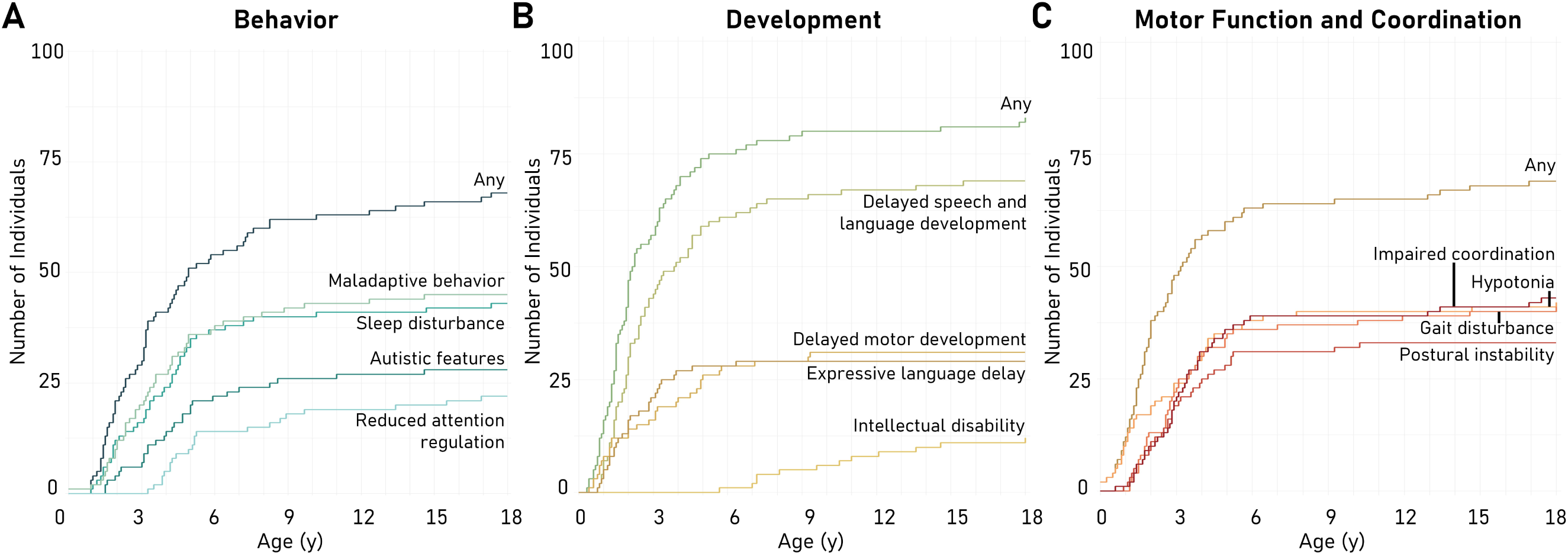
Behavioral, developmental, and motor coordination features of *SCN1A*-related disorders. Cumulative onset of behavioral (**A**), developmental (**B**), and motor coordination (**C**) phenotypes are shown over the first 18 years of life. The y-axis represents the cumulative number of individuals reported to have a particular phenotype. The x-axis represents age in years.

### EEG abnormalities in *SCN1A*-related disorders develop with age

Due to a high frequency of refractory epilepsy, individuals with *SCN1A*-related disorders often undergo frequent EEG monitoring. We leveraged reports of 569 EEGs performed on 96 individuals. 77% (440/569) of EEGs were performed before age four, during which time the average participant received approximately one EEG annually. 191 studies were normal, and many individuals (68/96) had both normal and abnormal EEGs (**Fig. 5A)**. Most EEGs were read as normal before nine months of age (81/146; **Fig. 5B**). EEG abnormalities before three months were limited to focal epileptiform discharges, almost exclusively occurring in the pGoF cohort due to their earlier onset of epilepsy (**Fig. 5C**). These abnormalities occurred in multiple brain regions—central, temporal, occipital, and parietal—and consisted of focal sharp waves, spikes, and spike-waves. Multifocal epileptiform discharges were also enriched in this population (6/9 pGoF, 14/87 pLoF; *P* = 0.002). With age, focal epileptiform discharges became more common in the pLoF cohort (50/89), consistent with their later onset of epilepsy. Across all participants, EEG anomalies included generalized epileptiform discharges (53/96), slowing (69/96), spike-wave complexes (48/96), and focal sharp waves (26/96). Overall, we leveraged clinical EEG data to demonstrate differences in EEG findings between pGoF and pLoF *SCN1A*-related disorders, particularly in early life.

**Figure 5.**
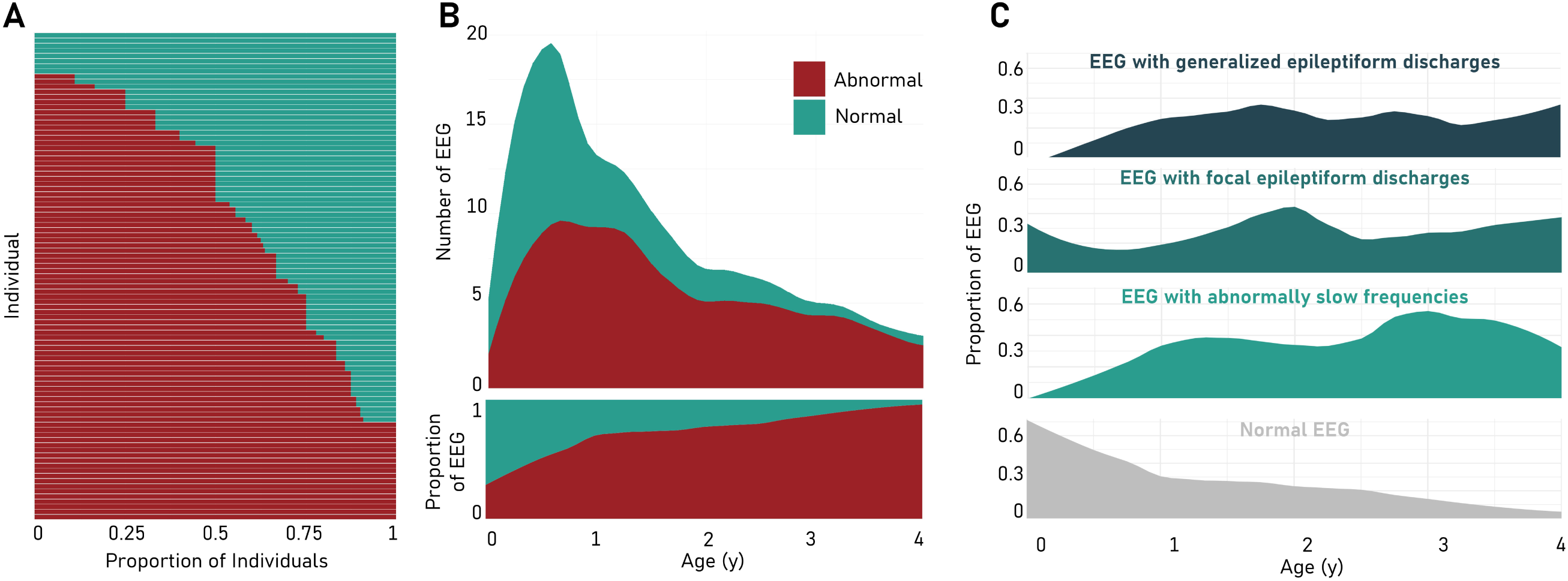
**EEG features of individuals with *SCN1A*-related disorders**. (**A**) Proportion of normal/abnormal EEGs (x-axis) per individual (y-axis). Color as indicated in legend. (**B**) Distribution of EEGs by age (x-axis) and by normal/abnormal status. Color as indicated in legend. (**C**) Smoothed frequency of different categories of EEG abnormality by age. X-axis represents age in years, and y-axis represents the proportion of EEGs with a particular abnormality. The proportion of normal EEGs are shown for reference in grey.

### The diagnosis of an *SCN1A*-related disorder is correlated with medication management changes in clinical practice

Seizure therapies, including medications, surgeries, and dietary approaches, were available for 95 patients, encompassing 554 therapies used over 1,405 patient years (**Fig. 6A**). Levetiracetam was the most frequently used medication, prescribed to 91% of participants (87/95), followed by valproic acid (69%, 66/95), clobazam (67%, 64/95), cannabidiol (57%, 54/95), and fenfluramine (53%, 50/95). The ketogenic diet was trialed in 35% of individuals (33/95). Surgical approaches, such as vagus nerve stimulation (11%, 10/95), were also recorded. Therapies were used for a median of 18 months. The ketogenic diet, clobazam, and cannabidiol were often used for extended periods (median 32, 26.5, and 23.5 months, respectively), whereas sodium channel blockers (SCBs)—contraindicated in pLoF *SCN1A*-related disorders—were used for shorter periods, such as oxcarbazepine, lacosamide, and lamotrigine (median 3, 3.5, and 5 months, respectively).

**Figure 6.**
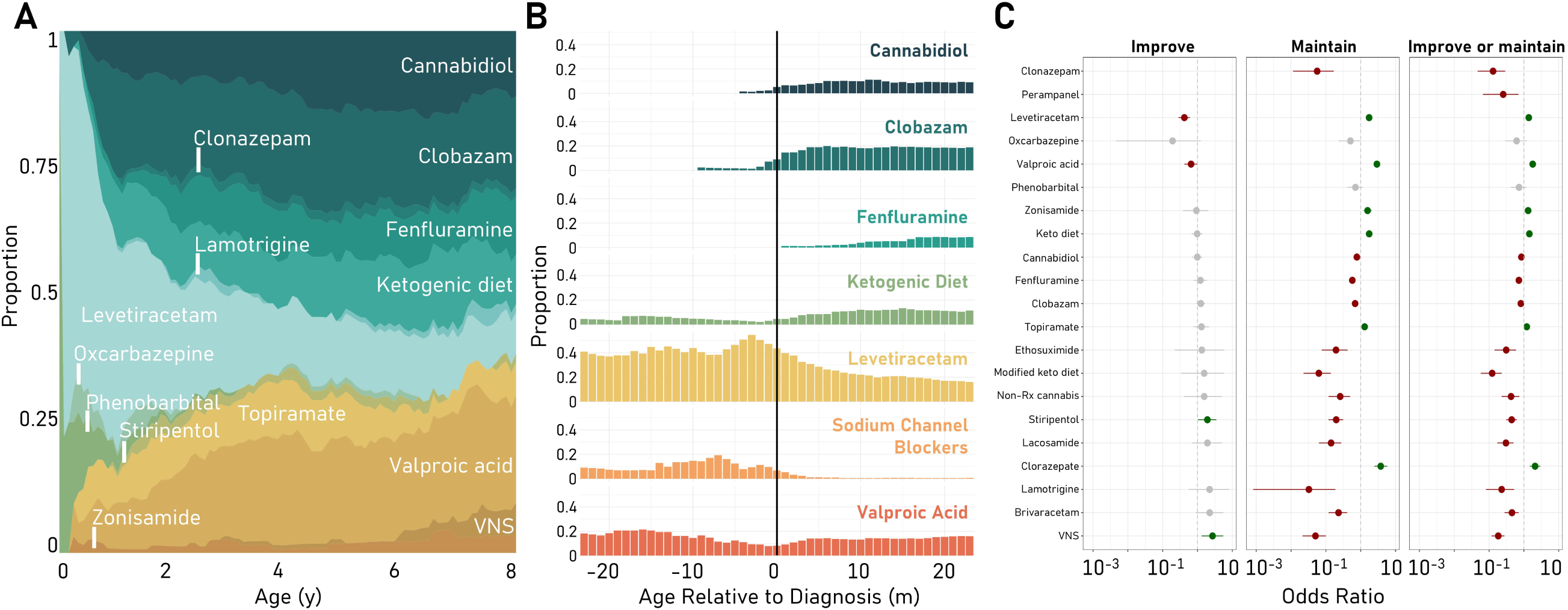
**Medication landscape and response to treatment**. (**A**) Anti-seizure therapies prescribed to individuals with predicted loss-of-function (pLoF) variants by age. Y-axis represents proportion of therapies used. X-axis represents age. (**B**) Proportion of individuals with pLoF variants on selected therapies before and after genetic diagnosis. Black vertical bar indicates the month of diagnosis. Y-axis represents the proportion of all active therapies, and x-axis represents age relative to month of genetic diagnosis. (**C**) Comparative efficacy of anti-seizure therapies at reducing seizure frequency (Improve), maintaining seizure freedom (Maintain), or both (Improve or maintain) in pLoF participants. Individual therapies are represented on the y-axis, and odds ratio is represented on the x-axis. Green circles represent an association with decreased seizure frequency or maintained seizure freedom (*P* < 0.05). Red circles represent an association with increased seizure frequency or failure to maintain seizure freedom (*P* < 0.05). Keto: ketogenic; non-Rx cannabis: cannabis obtained without medical prescription; VNS: vagus nerve stimulation.

Therapies also varied with age. In infancy, levetiracetam was used by most individuals with pLoF variants, a proportion that decreased sharply between 0–2 years of age due to the increased use of other therapies, including cannabidiol, clobazam, fenfluramine, the ketogenic diet, and valproic acid (**Fig. 6A**). Those with pGoF variants also had a high frequency of levetiracetam use in early life; however, the medications replacing its use were more often SCBs such as oxcarbazepine. In addition to age, the diagnosis of an *SCN1A*-related disorder coincided with changes in treatment. Diagnosis of a pLoF *SCN1A*-related disorder was often accompanied by the discontinuation of levetiracetam and SCBs and initiation of clobazam, cannabidiol, and fenfluramine. Genetic diagnosis did not affect all therapies; valproic acid and ketogenic diet use was similar before and after diagnosis (**Fig. 6B**). Among those with pGoF variants, levetiracetam was often discontinued, and clobazam initiated, following genetic diagnosis. Changes in SCBs—recommended for those with pGoF variants—were more ambiguous, with increased use of lacosamide but decreased oxcarbazepine use.

Treatment data were compared to seizure frequency to assess their comparative efficacies, a metric used in prior studies to identify therapies particularly efficacious in rare disease populations (**Fig. 6C**).^16^ For those with pLoF variants, stiripentol (*P* = 0.033) and vagus nerve stimulation (*P* = 0.003) were more associated with reduced seizure frequency. Levetiracetam (*P* < 0.001), valproic acid (*P* < 0.001), zonisamide (*P* < 0.001), the ketogenic diet (*P* < 0.001), topiramate (*P* = 0.006), and clorazepate (*P* < 0.001) were associated with an increased ability to maintain seizure freedom. The SCBs lacosamide (*P* < 0.001) and lamotrigine (*P* < 0.001) were associated with a lack of seizure freedom maintenance (**Fig. 6C**). Comparative effectiveness was not assessed for the pGoF cohort due to the small number of participants.

## Discussion

Here, we reconstructed the clinical trajectories of 100 individuals with *SCN1A*-related disorders across the lifespan, leveraging real-world data collected during routine clinical care. These data represent an abundant yet underused resource with the ability to complement the findings of prospective natural history studies. Standardized, retrospective, longitudinal disease trajectories were assembled for each participant—a novel paradigm as applied to *SCN1A*. Detailed information regarding the natural history of *SCN1A*-related disorders is essential to the provision of clinical care and anticipatory guidance, evaluation of approved therapies, and development of trial outcome measures in the era of precision medicine. We find that *SCN1A*-related disorders encompass distinct clinical syndromes, the diagnosis of which correlates with medication management changes, even though almost a quarter of diagnostic variants remain VUS.

### Seizure phenotypes in *SCN1A*-related disorders

The frequency and semiology of seizures were annotated from medical records encompassing 681 patient-years, adding granular, longitudinal data to existing knowledge of *SCN1A*-related disorders. The observed seizure landscape was dynamic, with a typical individual experiencing 17 changes in seizure frequency across the time assessed. Semiologies also evolved with age, with infancy being characterized by febrile, hemiclonic seizures, which were often superseded in childhood by focal impaired consciousness, atonic, and non-motor seizures. These evolutions in seizure type recapitulate the findings of prior publications and contribute to their temporal specificity.^34^

We then compared seizure phenotypes between individuals. A wide range of frequencies—from monthly to many daily—were observed. This range, particularly the prominence of monthly seizures, distinguishes *SCN1A*-related disorders from other genetic epilepsies, a comparison enabled by standardized seizure reconstruction. Epilepsy associated with *STXBP1*-related disorders, for example, is often characterized by seizures occurring at least daily, whereas seizure frequency in *SCN1A*-related disorders is typically monthly.^15^ This distinction persists even with other sodium channelopathies: Though a similar proportion of individuals with *SCN1A*- and *SCN8A*-related disorders will have seizures in a given month, most individuals with *SCN8A*-related disorders have daily seizures, a higher frequency than *SCN1A*-related epilepsies.^16^

We then compared epilepsy between provider-assigned clinical diagnoses, observing differences both in onset and frequency. Compared to Dravet syndrome, nd-DEE was associated with earlier onset and a four-fold increased prevalence of tonic seizures. GEFS+, conversely, was associated with a 27 times higher rate of simple febrile seizures—as opposed to prolonged, complex events—and relative absence of myoclonic seizures and convulsive status epilepticus. Notably, individuals diagnosed with atypical Dravet syndrome were highly similar to those with classical Dravet syndrome. This contributes to existing knowledge that, though each clinical diagnosis associated with *SCN1A* occurs on a spectrum, they each constitute a distinct disease entity without many true intermediate cases.^21^

The seizure trajectories assessed here reflect the natural history of *SCN1A*-related disorders under the best therapeutic strategies currently available. Notably, despite the multiple therapies trialed on and approved for individuals with Dravet syndrome, epilepsy in *SCN1A*-related disorders remains a challenge to manage and will benefit from the existence of true disease-modifying therapies. Many individuals in this study have ongoing seizures throughout childhood, including some with countable motor seizures at frequencies amenable for use as clinical trial outcomes. Conversely, many individuals with *SCN1A*-related disorders have seizures at monthly intervals. In fact, most participants had weekly or daily seizures for less than six months. Despite an ongoing seizure burden, these data demonstrate that many individuals with *SCN1A*-related disorders may not have frequent enough seizures to qualify for clinical trials using seizure frequency as a primary outcome measure. This suggests a need for alternative metrics, such as standardized developmental measures, for this population in a clinical trial setting.

### Developmental and co-morbid features of *SCN1A*-related disorders

We also analyzed non-seizure features of *SCN1A*-related disorders. Common groups of symptoms emerged, including developmental, behavioral, and motor coordination challenges. Developmentally, 83% of individuals were delayed, though early milestone acquisition was often within the range of normal. A formal autism diagnosis was made in 14/100 participants, a rate lower than the number of individuals displaying autistic features (28/100) as well as prior estimates of autism prevalence in *SCN1A*-related disorders, which range from 25% to 61% when assessed prospectively.^35,36^ This may reflect an underdiagnosis of autism among affected individuals, a phenomenon observed in other genetic neurodevelopmental disorders such as Fragile X syndrome, which may be influenced by difficulties diagnosing autism in individuals with co-occurring intellectual disability.^37,38^ It may also be the result of barriers to care, such as poor access to developmental pediatrics or the inaccessibility of mental health records for research to uphold patient privacy. Over 80% of the cohort, (91% excluding those with GEFS+) had cognitive differences, including developmental delay, intellectual disability, and significant impairments in communication. Developmental regression was not commonly observed, in broad agreement with data obtained from prospective natural history studies using standardized measures.^39^ Though early motor milestones were often achieved on time, motor coordination impairments emerged as a frequent challenge for participants, including difficulties relating to hypotonia, incoordination, and imbalance. With the exception of hypotonia, many of these symptoms emerged after the first year of life, reflecting the widening gap observed in prior studies between those affected by *SCN1A*-related disorders and their neurotypical peers over time.^39^ These results emphasize the burden of developmental and motor symptoms in *SCN1A*-related disorders while contributing to the conclusions of prospective studies suggesting that delayed symptom onset and developmental plateauing—not frank regression—characterize childhood with Dravet syndrome.^33^

### Anti-seizure therapies in *SCN1A*-related disorders

Comparing improvements in seizure frequency and maintenance of seizure freedom with epilepsy therapies permitted an assessment of the efficacy of these therapies in a routine clinical setting. Among the therapies trialed in individuals with pLoF variants, only stiripentol and vagus nerve stimulation were associated with a higher reduction in seizure frequency compared to other treatments. Other agents, including clorazepate, valproic acid, and the ketogenic diet were associated with improved maintenance of seizure freedom. Few therapies were broadly efficacious across the cohort, indicating the ongoing need for disease-modifying therapies. These data contrast with observations in clinical trials of anti-seizure medications. For example, fenfluramine did not stand out as a particularly efficacious medication for pLoF *SCN1A*-related disorders, despite being reported to reduce convulsive seizures in Dravet syndrome by 62% in a randomized control trial.^40^ Similarly, though a trial of cannabidiol showed a 50% reduction in seizures in 43% of treated individuals, it was not associated with a comparatively larger reduction in seizure frequency in this study. This distinction emphasizes that the real-world application of anti-seizure medications may not recapitulate the efficacy observed in trials conducted in highly controlled settings.

The genetic diagnosis of an *SCN1A*-related disorder correlated with changes in clinical management. Guidance on the avoidance of SCBs in individuals with pLoF variants appeared to be followed, with decreases in SCB use after genetic diagnosis. Recommendations to trial SCBs in those with pGoF variants, however, were not as clearly adhered to, which may be influenced by the recent description of this disease entity. The diagnosis of an *SCN1A*-related disorder coincided with the increased use of anti-seizure medications proposed by the international consensus on diagnosis and management of Dravet syndrome, such as clobazam, fenfluramine, and cannabidiol.^41^ Together, these observations demonstrate that the diagnosis of an *SCN1A*-related disorder is related to clinical management changes, even in the absence of a true disease-modifying therapy.

### Genetic variation associated with *SCN1A*-related disorders

Across the cohort, 80 *SCN1A* variants were observed. This is consistent with established knowledge suggesting that most patients with *SCN1A* haploinsufficiency harbor private variants. In fact, over 2,200 *SCN1A* variants have been classified as pathogenic or likely pathogenic in ClinVar. Half of individuals with pLoF variants had missense variants, raising potential challenges for variant interpretation. Applying ClinGen ESC-VCEP variant curation criteria, which are tailored to both the frequency and phenotypic specificity of *SCN1A*-related disorders, resulted in 61/80 variants—including 24/41 missense variants—meeting criteria for a pathogenic or likely pathogenic classification, leaving 19/80 (24%) VUS. Given that our study population is representative, this suggests that up to one in four individuals diagnosed with an *SCN1A*-related disorder harbors a VUS, even with a clinical diagnosis of Dravet syndrome. This highlights the ongoing need for improved variant classification measures, even among etiologies as well-researched as *SCN1A*. As precision therapies enter the clinical space, it will be of critical importance to determine whether a variant is diagnostic to discern the population of eligible recipients without misapplying therapies. Created by expert panels, ClinGen modifications to ACMG/AMP variant interpretation recommendations are an important step in this process, though not all VUS are resolved within this framework.^42–44^ Both functional studies of variants and additional genomic data on both unaffected and affected individuals will prove critical to this effort over time.

### Limitations

A primary limitation of this study is its restriction to the real-world data recorded during clinical care. These data are often heterogeneous and may be incomplete, as factors influencing medical decision-making may not align with those important to a study of natural history. Using a controlled biomedical dictionary, these challenges are at least partially addressed by enabling the comparison of features recorded at varying levels of detail. Additionally, by incorporating a larger cohort at relatively low cost, this methodology compensates for missing information in the EMR.

### Conclusions

In summary, real-world retrospective data was used to assess the natural history of *SCN1A*-related disorders, demonstrating a diversity of variants, clinical diagnoses, seizure trajectories, developmental outcomes, and medication response among affected individuals. Our results emphasize ongoing challenges in diagnostics and management of *SCN1A*-related disorders. Though improvements in genetic diagnostics and variant curation continue, 24% of identified variants remain variants of uncertain significance. Moreover, even with the approval of pharmacologic therapies trialed in individuals with *SCN1A*-related disorders, seizure burden in this population persists, with over 50% of individuals having seizures from age 1–5, and developmental features remain therapeutically unaddressed. This underscores the critical need for disease-modifying therapies in this population. Encouragingly, the real-world data compiled here contributes to a wealth of knowledge of the natural history of *SCN1A*-related disorders and demonstrates that their diagnosis is associated with recommended changes in clinical management, which are integral building blocks for the development and application of precision medicine approaches.

## Data availability

The data that support the findings of this study are available on request from the corresponding author. Data are not publicly available due to their containing information that could compromise the privacy of participant**s**.

## Data Availability

All data produced in the present study are available upon reasonable request to the authors.

## Acknowledgements

We would like to acknowledge the Dravet Syndrome Foundation and all participants for their ongoing support of this project.

## Funding

I.H. is supported by the National Institute for Neurological Disorders and Stroke (R01 NS131512 and NS127830). E.M.G. is supported by the National Institute for Neurological Disorders and Stroke (R01 NS110869 and NS137604). I.H. and E.M.G. are supported by the Dravet Syndrome Foundation (Research Grant).

## Competing interests

The authors declare no competing interests.

## Supplementary material

Supplementary material is available online.

## Notes

### Competing Interest Statement

The authors have declared no competing interest.

### Author Declarations

IRB of the Children's Hospital of Philadelphia gave ethical approval for this work.

